# Metabolomic and Immunologic Discriminators of MIS-C at Emergency Room Presentation

**DOI:** 10.1101/2024.01.11.24301110

**Authors:** Laura A. Vella, Amalia Z. Berna, Allison M. Blatz, Joey Logan, Priya Sharma, Yang Liu, Jonathan Tedesco, Cara Toland, Leena Babiker, Kathryn Hafertepe, Shane Kammerman, Josef Novacek, Elikplim Akaho, Alexander K. Gonzalez, Deanne Taylor, Caroline Diorio, Fran Balamuth, Hamid Bassiri, Audrey R. Odom John

## Abstract

Multisystem Inflammatory Syndrome in Childhood (MIS-C) follows SARS-CoV-2 infection and frequently leads to intensive care unit admission. The inability to rapidly discriminate MIS-C from similar febrile illnesses delays treatment and leads to misdiagnosis. To identify diagnostic discriminators at the time of emergency department presentation, we enrolled 104 children who met MIS-C screening criteria, 14 of whom were eventually diagnosed with MIS-C. Before treatment, we collected breath samples for volatiles and peripheral blood for measurement of plasma proteins and immune cell features. Clinical and laboratory features were used as inputs for a machine learning model to determine diagnostic importance. MIS-C was associated with significant changes in breath volatile organic compound (VOC) composition as well as increased plasma levels of secretory phospholipase A2 (PLA2G2A) and lipopolysaccharide binding protein (LBP). In an integrated model of all analytes, the proportion of TCRVβ21.3+ non-naive CD4 T cells expressing Ki-67 had a high sensitivity and specificity for MIS-C, with diagnostic accuracy further enhanced by low sodium and high PLA2G2A. We anticipate that accurate diagnosis will become increasingly difficult as MIS-C becomes less common. Clinical validation and application of this diagnostic model may improve outcomes in children presenting with multisystem febrile illnesses.

## INTRODUCTION

Multisystem Inflammatory Syndrome in Children (MIS-C) emerged in early 2020 as a sequela of SARS-CoV-2, typically appearing 4-6 weeks after first infection. The syndrome is characterized by fever, elevated markers of inflammation, and multi-organ system abnormalities. The most commonly used strategy for clinical screening follows the surveillance criteria established by the Centers for Disease Control (CDC), which have been iteratively updated to reflect the most current understanding of MIS-C^1^. Large clinical studies have suggested that the gastrointestinal and cardiovascular systems are most consistently involved, and more than 30% of patients develop shock^2^.

The surveillance criteria for MIS-C are broad and capture many non-MIS-C pediatric diagnoses. Indeed, overdiagnosis of MIS-C carried substantial risk for those misdiagnosed^3–5^. A study of 570 cases reported under the CDC surveillance guidelines identified 3 classes of cases, with the third class (198 patients) having clinical presentations that overlapped with other pediatric illnesses, particularly Kawasaki Disease^2^. Many other studies have identified clinical laboratory features that are increased in MIS-C, including elevated C-reactive protein (CRP), elevated absolute neutrophils, lymphopenia, elevated B-natriuretic peptide (BNP), and elevated D-dimer. However, many febrile illnesses are also associated with similar laboratory value changes, and to date, there is no single test that confirms or excludes the diagnosis of MIS-C in patients with a consistent clinical presentation.

Many research laboratory studies exploring unique features of MIS-C have focused on the immune system, observing derangements of both the innate and adaptive compartments^6,7^. Multiple groups have documented elevated plasma cytokine levels, increases in autoreactive antibodies, increases in T cell activation, and alterations in monocyte presentation machinery when children with MIS-C are compared to those with acute COVID-19, Kawasaki Disease, or other febrile illnesses^8–15^. The most consistent finding across studies has been an increase in the proportion of TCRVβ21.3+ T cells^8,15–19^. Still, even when these analytes are examined across large cohorts, there remains significant overlap between MIS-C and non-MIS-C controls, and no single immunologic feature fully discriminates MIS-C from other febrile illnesses. Additionally, many studies of immunologic parameters were performed on samples collected after therapeutic interventions with intravenous immunoglobulin, steroids, and/or cytokine-targeting biologics^14,20^ . Therefore, tools to assist in prompt, accurate MIS-C diagnosis at the time of initial clinical presentation are lacking.

In this study, we sought to enhance MIS-C diagnostic accuracy specifically at the time of initial emergency department evaluation. We enrolled children with clinical concern for MIS-C, operationalized as >3 days of fever and >2 organ system involvement or 1 organ system and known COVID-19 or exposure within the prior 4-6 weeks. Enrolled subjects provided breath and blood samples in addition to clinically indicated exam and laboratory studies. We enrolled more than 100 febrile children meeting MIS-C screening criteria and captured 14 children who received a final diagnosis of MIS-C. We also enrolled patients with acute SARS-CoV-2 who provided only breath samples. We then employed a machine learning model to provide a comprehensive and unbiased approach to determine feature importance in MIS-C diagnosis.

## RESULTS

### Machine learning to define MIS-C candidate predictors

We sought to identify clinical, metabolic, and immunologic features that distinguish children with and without MIS-C at the time of initial clinical presentation. Subjects were enrolled if they were under clinical suspicion for MIS-C in the emergency department of Children’s Hospital of Philadelphia prior to receipt of any immunomodulatory therapies. We defined clinical suspicion as use of a MIS-C clinical pathway and order set, which guided clinicians to consider MIS-C if children had >3 days of fever and >2 organ system involvement or 1 organ system and known COVID-19 or exposure within the prior 4-6 weeks. Our cohort consisted of 104 subjects, 14 of whom had MIS-C, enrolled between January 2021 and June 2022 (**Figure 1a**). Demographics and narrative summaries of the MIS-C and non-MIS-C cohorts are presented in **Table 1**. Children with and without MIS-C were broadly similar with respect to age, sex, and race characteristics. MIS-C patients were more likely to exhibit conjunctivitis (42.9% vs 21.1%, p=0.0003), compared to non-MIS-C cases.

**Figure 1:**
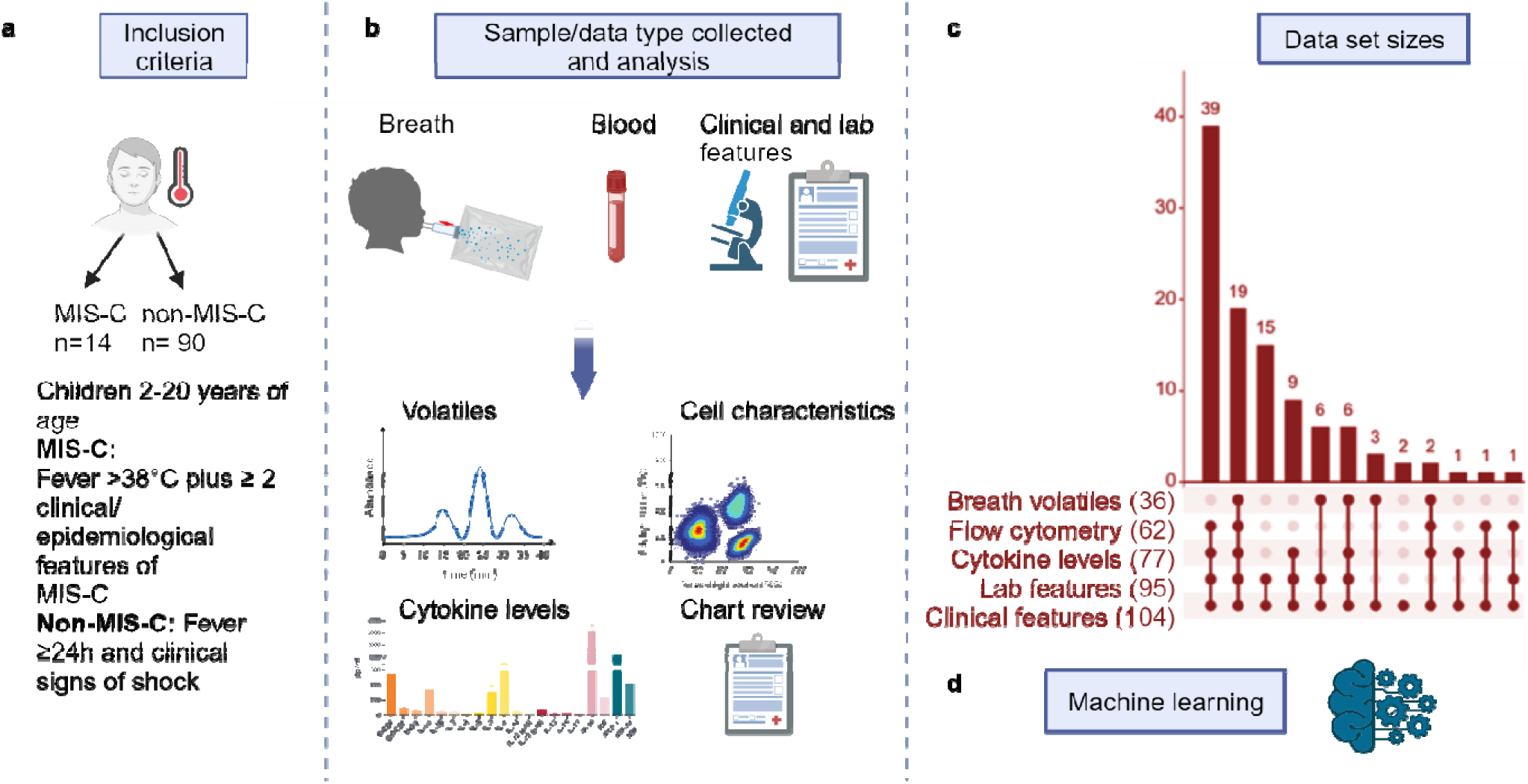
Schematic representation of subject cohorts and workflow, with number of individuals included in each analysis. **(a)** Inclusion criteria of MIS-C cohorts. **(b)** Breath samples, blood, laboratory, and clinical features were collected from enrolled children, and samples were analyzed using instruments detailed in methods section. **(c)** Number of samples available by various combination of assay (breath, flow cytometry, cytokines, laboratory and clinical features) is shown in vertical bars on the top of the diagram. The total number of patients analyzed/collected with each assay is indicated in parenthesis on the right of each assay. **(d)** Machine learning was applied to each data set and to the integrated data sets to find for features that best discriminate MIS-C from non-MIS-C cases.

**Table 1.** Demographic and clinical characteristics of enrolled patients.

| **Features** | **MIS-C**<br>**(n = 14)** | **Non-MIS-C**<br>**(n = 90)** | **p-value** |
| --- | --- | --- | --- |
| Age (years), median (IQR) | 7.9 (6.7-10.3) | 5.5 (6.7-10.3) | 0.35 |
| Race<br>White, n (%) | 10 (71.4) | 47 (52.2) | 0.39 |
| Sex<br>Female, n (%) | 6 (57.1) | 48 (46.7) | 0.57 |
| **Presenting symptoms, n (%)** |  |  |  |
| Fever (>38.0°C) | 14 (100) | 89 (98.9) | >0.99 |
| Diarrhea | 5 (35.7) | 36 (40.0) | >0.99 |
| Abdominal pain/emesis | 10 (71.4) | 53 (58.9) | 0.55 |
| Skin rash | 6 (42.9) | 33 (36.7) | 0.77 |
| Conjunctivitis | 6 (42.9) | 19 (21.1) | 0.0003 |
| Headache | 6 (42.9) | 37 (41.1) | >0.99 |
| Respiratory symptoms | 3 (21.4) | 40 (44.44) | 0.15 |
| COVID epidemiologic link | 5 (35.7) | 14 (15.6) | 0.12 |

Determination of MIS-C diagnosis was clinically decided by a multispecialty team of physicians and was additionally reviewed by two independent study team physicians (AMB, HB), with discordant reviews additionally analyzed by a third team physician (CD). Venous blood and exhaled breath were collected at the time of emergency department evaluation, and clinical and laboratory features from initial presentation were abstracted from the electronic medical record (EMR) (**Figure 1b**). As noted in **Figure 1c**, not all samples were available for each patient. To identify distinguishing features in each dataset, we built individual models for each data measurement class (clinical, laboratory, flow cytometry, cytokines, and breath metabolomics) using random forest classifier machine learning, as well as an integrated model using the top 8 features from each class (**Figure 1d**).

### Profound changes in clinical and laboratory features are observed in MIS-C

We first considered clinical and laboratory features that distinguish children with and without MIS-C (**Supplemental Table 1)**. Top 10 distinguishing features, as determined by random forest classification, are visualized in **Figure 2a**. Distinguishing clinical and demographic features of children with MIS-C included increased respiratory rate and maximum temperature (Tmax) as documented in the EMR. Furthermore, random forest classification identified race and ethnicity of children with MIS-C as important factors distinguishing MIS-C from non-MIS-C, which aligns with previously reported findings^21–23^. Top distinguishing laboratory features are visualized in **Figure 2b**, highlighting the marked differences in multiple laboratory parameters in children with MIS-C, including serum sodium, inflammatory markers (including C-reactive protein [CRP] and erythrocyte sedimentation rate [ESR]) and multiple hematological parameters. Of particular note, Tmax, CRP, and serum sodium were each statistically significantly different in children with MIS-C compared to febrile children (**Figure 2c, d, e**).

**Figure 2.**
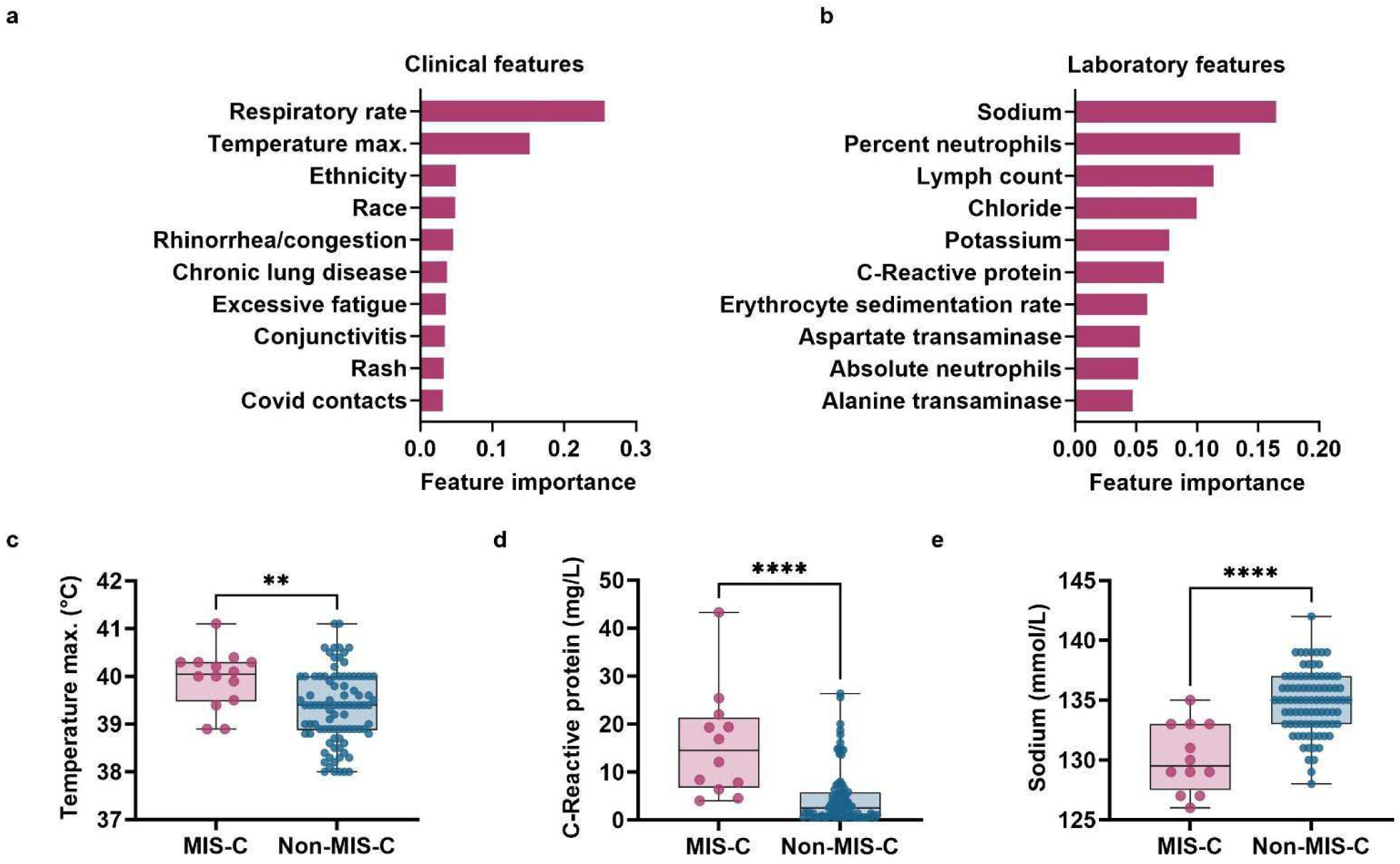
Alteration of clinical and laboratory features in MIS-C. Top 10 distinguishing clinical **(a)** and laboratory **(b)** features obtained after machine learning. **(c, d, e)** Box plots of maximum temperature, CRP and sodium levels respectively, in MIS-C and non-MIS-C cohorts. Whiskers represent maximum and minimum. Bars represent the median of each group. Significance was determined by unpaired Wilcoxon tests between clinical categories, indicated by ** p<0.01, **** p<0.0001.

### The breath composition of MIS-C patients is characterized by enrichment of terpenes

There is increasing interest in use of breath volatile biomarkers for diagnosis of a number of infectious and inflammatory syndromes [reviewed in^24^], including recent FDA approval of a breath-based diagnostic test for SARS-CoV-2^25,26^. We have previously established reproducible breath biomarkers of pediatric SARS-CoV-2 infection^27^. Since breath-based diagnostics have potential as low-cost point-of-care diagnostics suitable for low resource settings, we sought to evaluate whether the composition of exhaled breath was distinct in children with MIS-C, as compared to febrile controls. We found 29 distinct molecules in the breath of children with MIS-C. The overall breath volatile profile of children with MIS-C was highly distinct from that of children suspected of MIS-C (**Figure 3a**). We have previously identified characteristic aldehydes that are enriched in the breath volatile composition of children with SARS-CoV-2^27^. To evaluate whether MIS-C breath signatures were distinct from those of acute SARS-CoV-2 infection, we interrogated breath samples from children with SARS-CoV-2 (n=36) for the presence of the characteristic MIS-C volatiles. Breath samples from children with SARS-CoV-2 infection were collected and analyzed using identical methodology^27^. We find that children with MIS-C had a highly unique breath metabolite profile (**Figure 3a**). Interestingly, many of the most distinguishing breath volatiles (including terpinene, limonene, myrcene, carveol, and others) belong to a class of compounds called terpenes. These compounds were elevated in the breath of MIS-C patients compared to those febrile non-MIS-C and SARS-CoV-2 positive patients. Terpenes are a class of biomolecules that are typically associated with plants and microbial metabolism^28,29^. While humans produce the biosynthetic precursors to terpenes, they lack the terpene synthase enzymes that cyclize these metabolites to produce the final products^30^. Next, we used random forest analysis to evaluate the most influencing volatiles (**Figure 3b**). We find that elevated breath terpenes, especially γ-terpinene, to be highly diagnostic for MIS-C (**Figure 3c**). Levels of γ-terpinene were both sensitive and specific for diagnosis of MIS-C versus non-MIS-C in children with clinical symptoms prompting emergency room evaluation, with a receiver operating curve of 0.91 for this single volatile analyte (**Figure 3d**).

**Figure 3.**
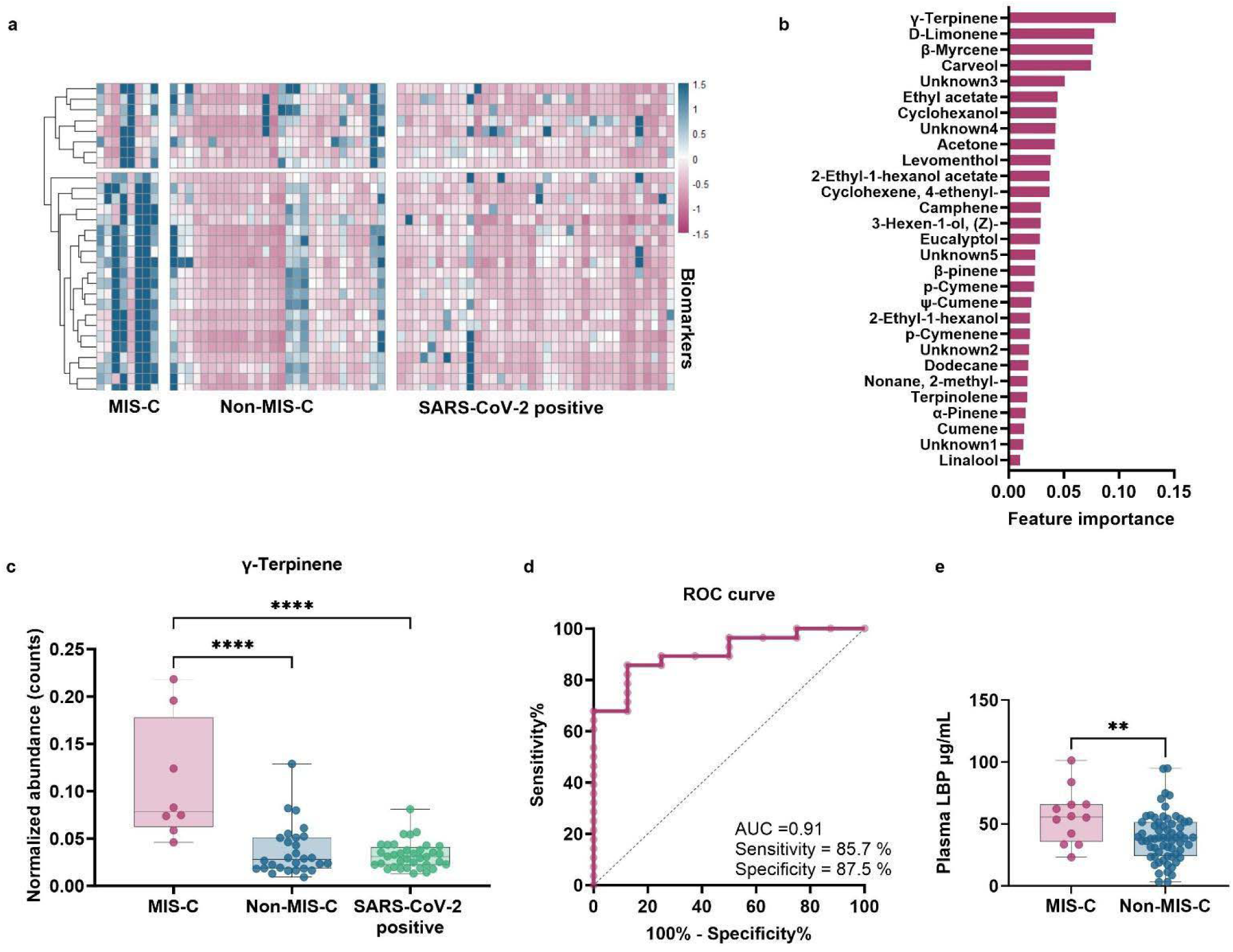
Untargeted analysis of breath volatiles reveals the change of 29 unique compounds in the breath of MIS-C patients. **(a)** Heat map visualizing abundance of 29 unique breath compounds (presented as z-scores) in MIS-C and non-MIS-C cohorts, and in SARS-CoV-2-infected pediatric subjects. Identity of 29 compounds can be found in **Supplemental Table 2**. **(b)** Feature importance plot for all unique volatiles. **(c)** Box plot of ү-terpinene. Whiskers represent maximum and minimum, and bar represents the median of each group. Significance determined by multicomparisons using Bonferroni test between three clinical categories, indicated by **** p<0.0001. Lack of notation for specified comparisons indicates no statistical significance. **(d)** Receiver operator characteristics (ROC) curve for the abundance of ү-terpinene. Dotted line indicates expected results if predictive power is no better than random chance. **(e)** Plasma lipopolysaccharide-binding protein (LBP). Whiskers represent maximum and minimum, and bar represents the median of each group. Significance determined by unpaired Wilcoxon test between MIS-C and non-MIS-C categories, indicated by ** p<0.01.

To evaluate whether elevated breath terpenes were associated with changes in intestinal barrier function, we evaluated the serum levels of lipopolysaccharide-binding protein (LBP), a biomarker of intestinal integrity. We find that LBP levels are elevated in MIS-C patients compared to non-MIS-C patients (**Figure 3e**) suggesting that the prominent GI symptoms in this cohort are correlated with a defect in intestinal integrity.

### Distinct cytokine and plasma protein profiles of MIS-C

We previously reported changes in plasma analytes in patients diagnosed with MIS-C (soluble proteins including cytokines and non-cytokines)^11,31^. These prior analyses were performed in hospitalized patients, some of whom had received anti-inflammatory therapies such as intravenous immunoglobulin (IVIG) and corticosteroids. These analyses identified elevations in plasma interleukin 10 (IL-10), tumor necrosis factor alpha (TNF-α), as well as increases in plasma gamma interferon (IFN-γ) in patients with milder presentations of MIS-C. As prior results may have been affected by immunomodulation, in the current study we used plasma from subjects collected at the time of evaluation in the emergency department (and prior to receipt of any immunomodulatory medications). Plasma cytokines were quantified using Meso Scale Discovery multiplexed assays, and additional soluble proteins [PLA2G2A, the complement membrane attack complex (sC5b9) and lipopolysaccharide binding protein (LBP)] were quantified by ELISA (**Supplemental Table 1**). We found that the plasma of subjects with MIS-C had elevated levels of TNF-α as compared to non-MIS-C patients presenting with fever, but that the elevated levels of IL-10 previously reported did not distinguish MIS-C patients from other febrile control patients (**Figures 4a** and **4b**).

**Figure 4.**
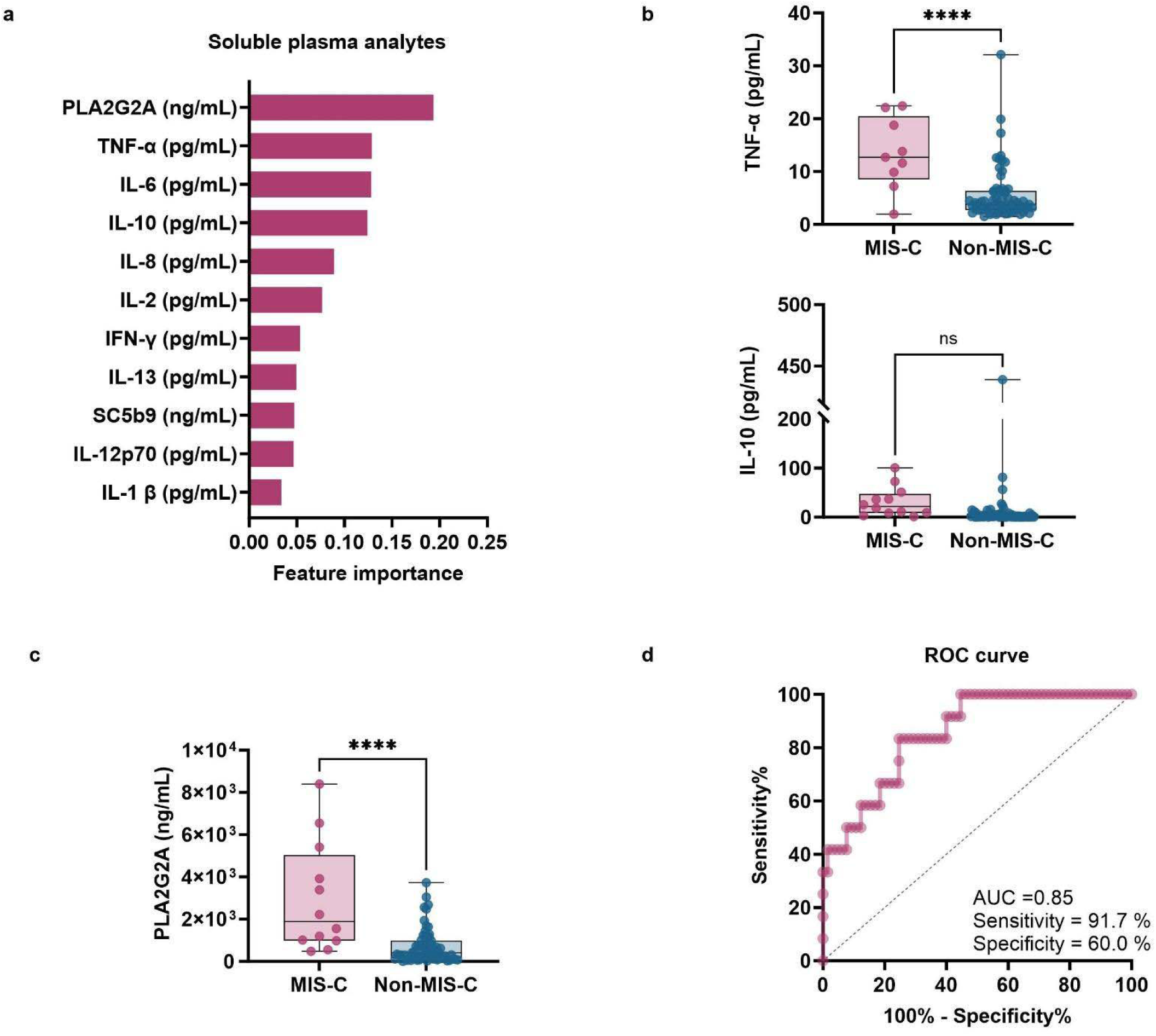
Cytokine signatures are important markers of MIS-C. **(a)** Feature importance plot for all cytokine markers. **(b)** Box plots of the two most differentiating cytokines (IL-10, TNF-alpha) in the plasma of MIS-C patients and **(c)** PLA2G2A. Whiskers represent maximums and minimums and bars represent the median of each group. Significance determined by unpaired Wilcoxon test between MIS-C and non-MIS-C categories, indicated by **** p<0.0001; ns indicates no statistical significance. **(d)** Receiver operator characteristics (ROC) curve for the abundance of PLA2G2A. Dotted line indicates expected results if predictive power is no better than random chance.

We had previously identified significant elevations in phospholipase A2 Group IIA (PLA2G2A) in patients with MIS-C^32^. PLA2G2A modulates membrane phospholipid composition and its expression is upregulated by inflammatory stimuli, as well as pro-inflammatory cytokines such as interleukin 1-beta (IL-1β) and interferon gamma (IFN-γ). In the current cohort, we find that the plasma of MIS-C patients contains significant elevations of PLA2G2A in comparison to that of non-MIS-C subjects (**Figure 4c**), even prior to receipt of immunomodulation. In fact, PLA2G2A levels represent one of the features most likely to distinguish MIS-C from other febrile conditions of children (**Figure 4a**), with an estimated sensitivity of 91.7% based on our receiver operator characteristic analysis (**Figure 4d**).

We similarly observe elevations in the lipopolysaccharide binding protein (LBP) in the plasma of children with MIS-C (**Figure 3e**). At low levels, LBP serves to bind lipopolysaccharides (LPS; endotoxins found on the outer membranes of Gram negative bacteria) and to present it to Toll-like receptor 4 (TLR4) and CD14 expressed by monocytes and macrophages^33^; in turn, these cells then secrete IL-1 and TNF-α. At higher levels, LBP may bind excess LPS and prevent overactivation of monocytes and macrophages in sepsis states^34,35^. LBP is produced by hepatocytes and intestinal epithelial cells^36^, and its levels rise rapidly in response to exposure to IL-1 and IL-6^37^.

### Immunologic markers define proliferation of TCRV**β**21.3+ T cells

One of the most consistent findings across studies of the immune state in MIS-C is the expansion of T cells that utilize the T cell receptor Vβ segment 21.3 or increases in the corresponding gene TRBV11-2, first identified by Porritt *et al*^17^. Several studies showed that approximately half of patients clinically diagnosed with MIS-C have evidence of TCRVβ21.3/TRBV11-2 expansion^8,15,17,19^. To determine whether a deeper focus on the phenotype of TCRVβ21.3 T cells and broader measures of immune state could improve diagnostic accuracy in MIS-C, we performed high-parameter spectral cytometry on cryopreserved peripheral blood mononuclear cells (PBMC) from the emergency room blood samples. We then built a random forest model with more than 90 immune features of interest, including T cell differentiation states defined as previously described^13^ and 20 features related to TCRVβ21.3 positive and negative cells (**Supplemental Table 1**). The proportion of Ki67+TCRVβ21.3+ non-naïve (nn) CD4 T cells was determined to be the top discriminator of MIS-C from controls at emergency room presentation, followed closely by exhaustion-like PD1+CD39+TCRVβ21.3+ non-naive (nn) CD4 T cells (**Figure 5a**). The percent Ki-67+ TCRVβ21.3+ nnCD4 T cells demonstrated a specificity of 87% and a sensitivity of 89% against the final clinical diagnosis (**Figure 5b**). When viewed as an individual flow cytometric measure, the difference in median values between MIS-C and non-MIS-C was approximately 10-fold (**Figures 5c**). We additionally considered that a metric using Ki-67 expression may falsely diagnose MIS-C when T cell proliferation is high across multiple T cells utilizing a diverse pool of Vβ gene segments. We therefore normalized the Ki-67 parameter within each patient using a ratio against an internal control of Ki-67 expression in TCRVβ21.3-nnCD4 T cells (**Figure 5d**). Indeed, the finding was robust to correction for total T cell proliferation, and the ratio even outperformed the frequency metric in sensitivity and specificity (**Figure 5e**), suggesting that deeper analyses of TCRVβ21.3+ T cells can be sensitive and specific markers of MIS-C.

**Figure 5.**
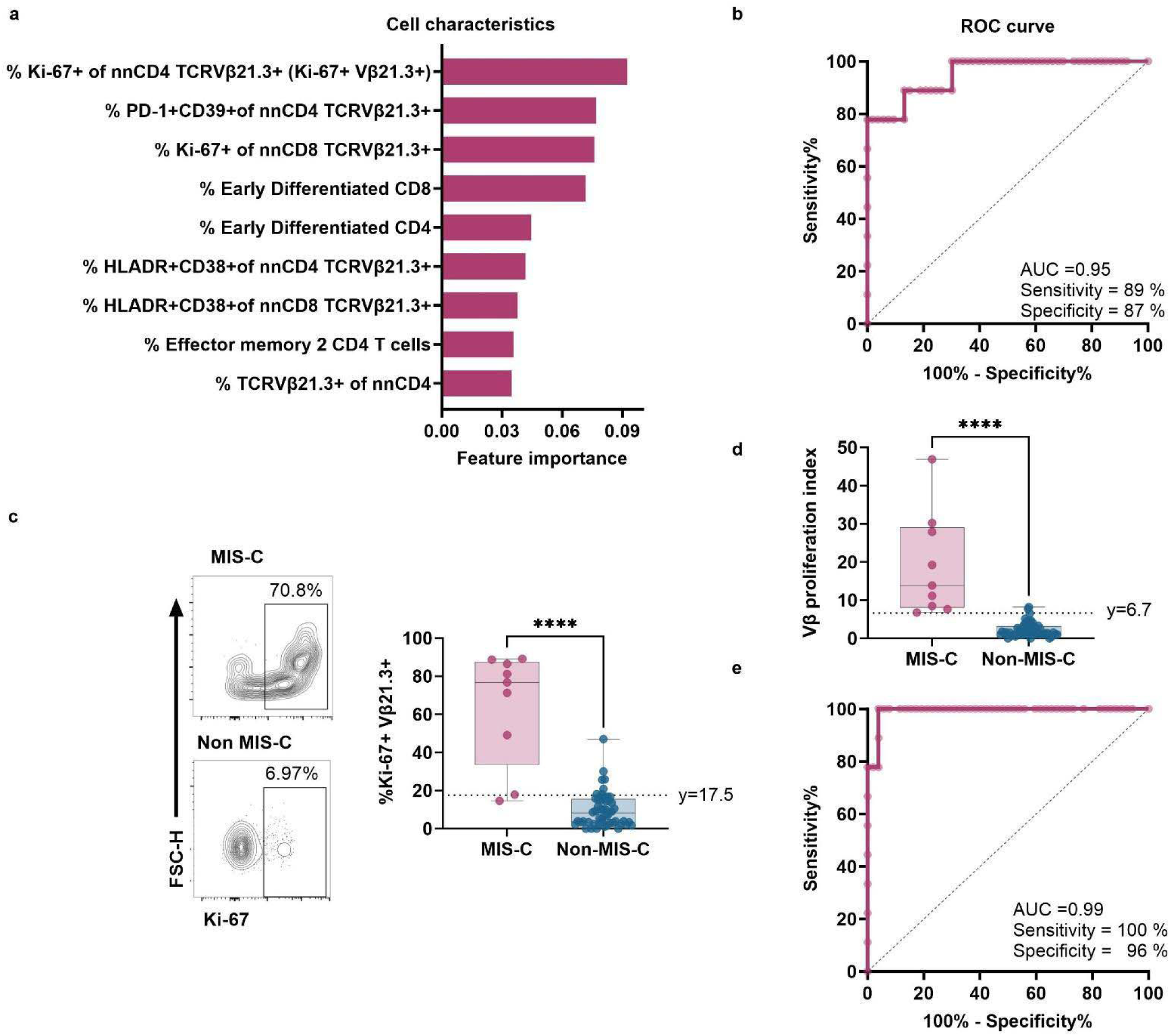
Co-expression of Ki-67 in TCRVβ21.3+ CD4 T cells enhances diagnostic accuracy in MIS-C. **(a)** Feature importance plot of flow cytometric measures. **(b)** Receiver operator characteristics (ROC) curve for the frequency of Ki-67 expression in non-naive (nn) CD4 TCRVβ21.3 T cells. Dotted line indicates expected results if predictive power were no better than random chance. Early differentiated T cells are CD27+CD45RA-. Effector memory 2 cells are defined as CD45RA-CD27-. **(c)** Frequency of identified feature in MIS-C and non-MIS-C with example flow cytometry plots from samples at the median of each group. **(d)** VL proliferation index. (e) ROC curve for VL proliferation index. Whiskers represent maximums and minimums, and bars represent the median of each group. Significance determined by unpaired Wilcoxon test between MIS-C and non-MIS-C categories, indicated by **** p<0.0001. FSC-H, forward scatter height.

### Integrated model identifies novel diagnostic strategy for MIS-C

Feature importance analysis based on random forest classification that includes all assays was performed. Since inclusion of breath volatiles measurements to the integrated model decreased the power of our analysis (due to small sample sizes), these were removed from the model. In total, 58 samples were used for the integrated model, and it was found that Ki-67+ TCRVβ21.3+ nnCD4 T cells, CRP, sodium, and PLA2G2A were the most important parameters distinguishing MIS-C from non-MIS-C (**Figure 6a**).

**Figure 6.**
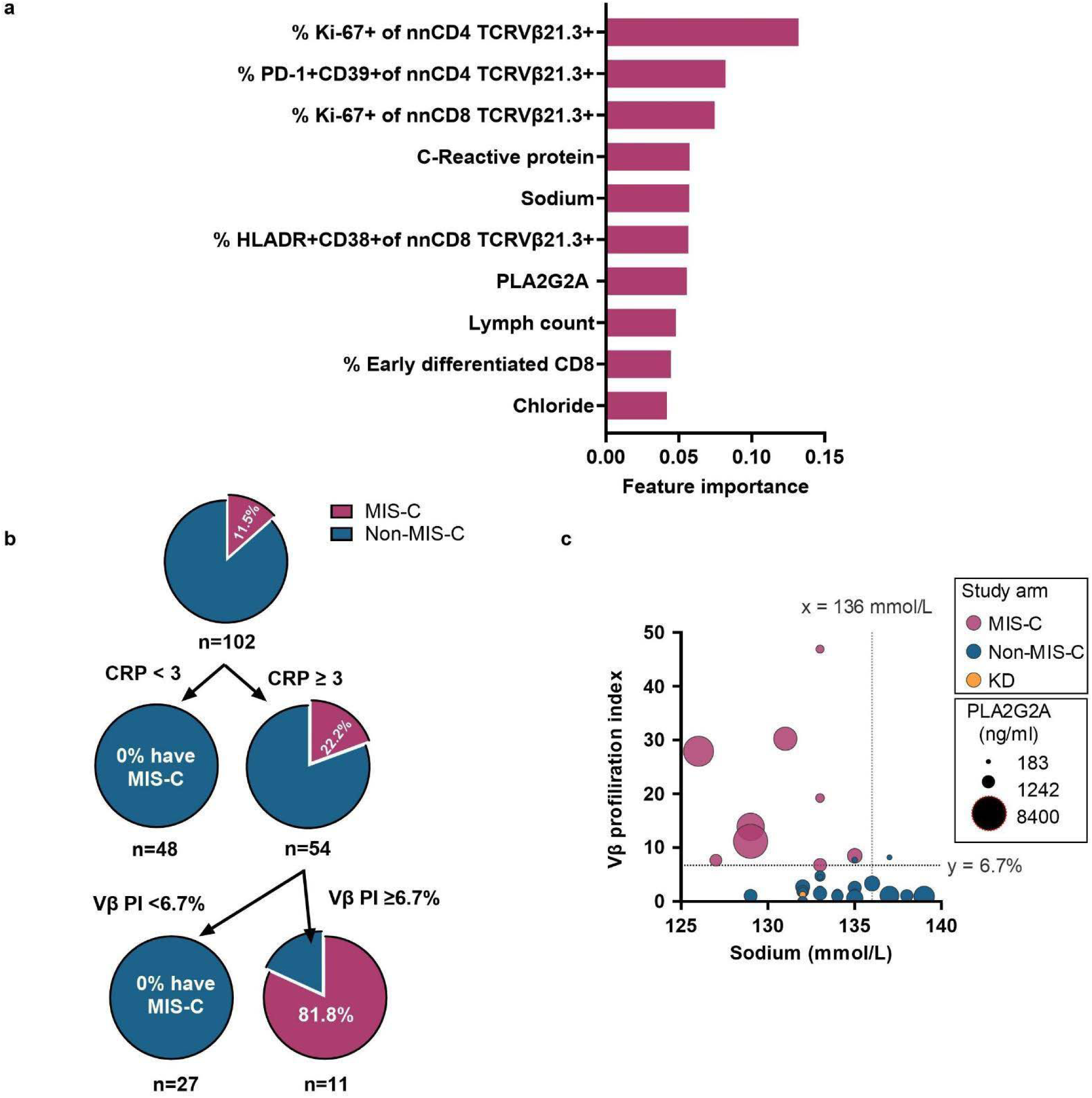
**Integrated model and MIS-C diagnostic strategy. (a**) Feature importance plot for top 10 markers after integrated model. **(b)** Diagnostic strategy using CRP and VL proliferation index (VL PI) **(c)** Correlation between blood sodium levels, VL proliferation index and PLA2G2A. Size of dots represents concentration range of PLA2G2A and colors represent MIS-C, non-MIS-C, and Kawasaki disease patients.

C-reactive protein (CRP) ≥3.0 mg/dL is one of the recommended criteria used for identification and classification of cases of MIS-C . In our study we used the CRP threshold on 102 subjects (12 MIS-C positive cases), resulting in 54 subjects with CRP ≥3.0 mg/dL. All MIS-C positive cases fell within this category; however, a large number of non-MIS-C patients also had CRP elevations (**Figure 6b**). To stratify patients with characteristic immune profiles associated with MIS-C, we developed a “Vβ proliferation index” (ratio of Ki-67+TCRVβ21.3+ to Ki-67+TCRVβ21.3-nn CD4 T cells). Applying a threshold of 6.7% on the “Vβ proliferation index,” we identified 11 patients above this threshold, which included all patients diagnosed with MIS-C and only 2 subjects who did not receive that diagnosis (**Figure 6b**).

Additionally, we looked for correlations among the ratio of Ki-67+TCRVb21.3 +/-, blood sodium and PLA2G2A and found that all MIS-C positive subjects showed low blood sodium (<136 mmol/L), Ki-67+ TCRVβ21.3 +/- >6.7% and elevated PLA2G2A (>660 ng/ml) (**Figure 6c**).

## DISCUSSION

The CDC case definition for MIS-C remains a diagnosis of exclusion, in which MIS-C is rejected if there are “other more likely alternative diagnoses,” yet the clinical and laboratory features of MIS-C largely overlap with those of many other childhood infectious and/or inflammatory conditions. This has led to many anecdotal and published reports regarding MIS-C misdiagnosis^39,40^. To address this diagnostic uncertainty during initial presentation in the acute care setting, we used a machine learning approach to identify key clinical and laboratory parameters which, when combined, could provide significant power in discriminating MIS-C in children presenting with fever and clinical suspicion for MIS-C. Our study comprises one of the largest prospectively collected cohorts of untreated MIS-C subject samples. Importantly, our study compares biomarkers in untreated MIS-C patients against the most challenging and relevant control group: children without MIS-C whose presentations were similar enough to prompt clinical suspicion and laboratory evaluation for the possibility of MIS-C. We have identified a commonly available clinical laboratory feature of inflammation (CRP), which when combined with a “Vβ proliferation index,” correctly identified 100% of patients with MIS-C presenting to a free-standing tertiary care urban children’s hospital over an 18-month period.

Our data inform the potential clinical value of a “Vβ proliferation index” (ratio of Ki-67+TCRVβ21.3+ to Ki- 67+TCRVb21.3- non-naive CD4 T cells) to improve the diagnostic evaluation of suspected MIS-C. While expansion of TCRVβ21.3+ cells amongst total T cells has been previously observed in MIS-C (reviewed in^7^), we find that sensitivity is enhanced by focusing on non-naive CD4 T cells and by measuring activation-associated features of TCRVβ21.3+CD4+ T cells instead of TCRVβ21.3 frequency alone. The validity of this approach is mirrored in the flow cytometric TCRVβ21.3 data in MIS-C published by Moreews *et al* in 2021^8^. While the numbers were limited for in-depth flow cytometric analysis of TCRVβ21.3+ cells, their data clearly showed the largest separation between cases and controls in the co-expression of activation markers HLA-DR and CD38 amongst TCRVβ21.3+ and not TCRVβ21.3-CD4 T cells, with the same analysis in CD8 T cells demonstrating comparatively less separation. However, in their study, only 1 patient diagnosed with MIS-C who did not have detectable TCRVβ21.3 expansion demonstrated increased activation of TCRVb21.3+ CD4 T cells. We propose that focusing on non-naive CD4s and using Ki-67 has improved sensitivity for detection of MIS-C.

Our results also provide the first profile of the breath composition of children with MIS-C. This study revealed a distinctive breath profile in children with MIS-C, setting it apart from febrile controls with similar clinical features and from those with acute SARS-CoV-2 infection. Furthermore, our analyses uncovered a significant enrichment of terpenes, particularly gamma-terpinene, in the breath of MIS-C patients. As humans do not synthesize terpenes directly this unique breath signature raises intriguing questions about their possible origin in the diet or microbiome, especially given the increase in serum LBP levels, which suggests a loss of intestinal barrier dysfunction. Terpenes have previously been found more abundant in the feces of patients with diarrhea-predominant IBS compared to healthy controls^41^.

There are a few limitations in our study. Reflecting the rarity of MIS-C, our single center cohort contains small numbers of subjects with this diagnosis; similarly, our cohort contains few subjects with alternative diagnoses that have substantial clinical overlap with MIS-C, such as Kawasaki disease or scrub typhus infection. Despite the uncertainty associated with the diagnosis of MIS-C, for which there is no gold standard, our machine learning approach clearly highlighted diagnostic biomarkers with strong discriminating power. Prospective studies will be required to validate the diagnostic accuracy of the Vβ proliferation index. Although validation studies will be challenging due to the reduced incidence of MIS-C, our data suggest that such validation is worthwhile to determine whether the use of flow cytometric assessment of TCRVβ21.3+ is a simple potential discriminator when children with clinical features of MIS-C and elevated CRP are assessed in the emergency room setting. Fast and accurate diagnostic tools are required both to avoid misdiagnosis and to ensure timely administration of immunomodulatory treatment to avoid intensive care unit admission and sequelae of delayed therapy^42^.

## Data Availability

All data produced in the present study are either contained within the manuscript or available upon reasonable request to the authors

## ACKNOWLEDGEMENTS

We thank the children and families who participated in this study, along with the staff of the Children’s Hospital of Philadelphia for their support. We thank Hannah Edelman and Madison Reilly for assistance with subject enrollment and sample collection. Research reported in this publication was supported by NIH T32 GM075766 (AMB), NIH/NIAID K08 AI136660 (LV), NIH/NIAID R21AI154370 (AOJ), NIH/NICHD R01HD109963 (AOJ), and NIH/NICHD R61/33HD105594 (AOJ, HB, LV, FB, AB). Dr. John is an Investigator in the Pathogenesis of Infectious Diseases of the Burroughs Welcome Fund. Dr. Vella’s work was supported by the Doris Duke Foundation Grant #: 2021190. CJD is supported by a Canadian Institute for Health Research (CIHR) Fellows Award and 5K12CA076931-24. The funding sources had no role in the design, conduct, or analyses of this study.

## METHODS

### Study Approval and Enrollment

Prior to enrollment, the study was approved by the Children’s Hospital of Philadelphia (CHOP) Human Research Ethics Committee (IRB 20-018139 and IRB 20-017503) and by the CHOP Institutional Biosafety Committee (IBC 19-000145) for handling of human samples potentially containing SARS-CoV-2. Participants’ or their legal representatives’ consent to take part in the present study.

We enrolled febrile children 2-20 years of age presenting for care in the Children’s Hospital of Philadelphia Emergency Department (ED) from January 2021 to June 2022, with symptoms consistent with MIS-C. This timeframe encompasses circulating SARS-CoV-2 lineages preceding Omicron. Clinical concern for MIS-C was defined as fever >38.0°C for ≥ 3 days, plus ≥ 2 clinical/epidemiologic features suggestive of MIS-C (rash, gastrointestinal symptoms, mucosal changes, extremity changes, neurological symptoms, lymphadenopathy, epidemiological link to COVID-19). In addition, patients were enrolled with fever ≥24h and clinical signs of shock, even in the absence of additional clinical/epidemiological symptoms. Children that were eligible to participate had the choice to provide breath and blood samples. All patients were enrolled prior to the receipt of immunomodulatory agents, including intravenous immunoglobulin and steroids.

For SARS-CoV-2-infected subjects and breath samples only, we collected samples from children (4-18 years of age) admitted to the Extended Care Unit of the Emergency Department of the Children’s Hospital of Philadelphia, who had been diagnosed as SARS-CoV-2 positive. COVID-19 test results included the following: i) tests from other clinical institutions, ii) RDT-home antigen tests; or iii) RDT performed by CHOP ED study members as a study procedure.

Subjects were excluded from all studies for the following reasons: oxygen supplementation within 3 hours of breath sample collection, pre-existing neurological disorder, Wilson’s disease, or diabetes mellitus requiring treatment with insulin. Specific for breath collection, children <4 year of age, with altered metal status, or who are unable to cooperate with voluntary collection were also excluded from the study.

### Data collection

Clinical data was abstracted from electronic patient charts on to standardized case report forms created using the Research Electronic Data Capture database (REDCap version 13.11.4; Vanderbilt University, Nashville TN USA). Data were abstracted by a clinician or clinical research assistant. Laboratory data was extracted from Electronic Health Record. All data elements were validated by a physician. Data collected included demographics, comorbid conditions, sources of coinfection, gastrointestinal symptoms, most extreme laboratory values for laboratory studies of interest and laboratory indicators of organ dysfunction or inflammation. The IDs used in Supplemental Table 1 are not known to anyone outside the research group.

### Breath Collection

Breath collection was performed as previously described^43,44^. In brief, subjects exhaled through a disposable cardboard mouthpiece connected to a chamber. The chamber was then attached using tubing to a 3-L SamplePro FlexFilm sample bag (SKC Inc, Pennsylvania; Supplementary Figure 1). Subjects were coached to take a few deep breaths, place the cardboard tube between the lips, and exhale completely. Neither a nose clip nor VOC filter were used. Breath from the bags was transferred to a sorbent tube as previously described^43^. Briefly, 1 L of the breath sample was transferred to sorbent tube at 200 mL minL1 using an electric pump, so that all tubes had consistently the same volume. Three-bed Universal sorbent tubes containing Tenax, Carbograph, and Carboxen were used (Markes International Limited, UK). For each participant, ambient air samples and breath samples were collected from the same room at the time of sample collection. Samples were stored at 4°C until the time of analysis, and all analyses were completed within 2 weeks of sample collection. Samples were analyzed using thermal desorption and GCxGC BenchTOF-MS (SepSolve Analytical, UK), as described below.

### Blood Collection and Processing

Venous blood was collected into both sodium and lithium heparin tubes. Sodium heparin tubes were maintained at room temperature until time of processing, typically within 12-18hrs of collection. Blood was spun for plasma collection and then processed for peripheral blood mononuclear cells (PBMC) using Sepmate isolation kits. PBMC were cryopreserved in 10% DMSO in FBS for later use in flow cytometric assays. Lithium heparin tubes were maintained at 4°C until processing. Blood was spun and plasma was cryopreserved before use in cytokine assays.

### Flow Cytometry

Samples were thawed in batches that included MIS-C cases and controls. Samples were stained with a viability dye followed by treatment with human Fc block (BD Biosciences #564219). Samples were then stained for chemokine receptors at 37°C. Although data not included, cells underwent incubation with B cell probes for SARS-CoV-2 proteins prior to proceeding to staining with the remaining surface protein-specific antibodies. Samples were subsequently permeabilized with Foxp3 / transcription factor fixation/permeabilization concentrate and diluent (Invitrogen Cat# 00-5521-00) before proceeding to intracellular protein stains (**Supplemental Table 3**). Stained samples were fixed with 1% paraformaldehyde before acquiring on a Cytek Aurora cytometer. Demultiplexed data were gated and analyzed using FlowJo (TreeStar).

### Soluble Plasma Analyte Measurements

#### Proinflammatory cytokine profiling

Ten proinflammatory cytokines were measured (IFN-γ, IL-1β, IL-2, IL-4, IL-6, IL-8, IL-10, IL-12p70, IL-13, and TNF-α) using V-Plex Pro-inflammatory Panel 1 Human Kits (catalog #K15049D; Meso Scale Diagnostics, Rockville, MD). All assays were performed in duplicate, according to the manufacturer’s protocol. Assays were read and analyzed on a QuickPlex SQ120 (Meso Scale Diagnostics).

#### sC5b9 assay

Plasma samples were assayed for sC5b9 by using a human C5b9 enzyme-linked immunosorbent assay (ELISA) set (#558315; BD Biosciences, San Jose, CA) according to the manufacturer’s protocols. All samples were assayed in triplicate. Standard curves were used to derive sC5b9 levels from best fit curves.

#### PLA2G2A assay

Plasma samples were assayed for PLA2G2A by using a human PLA2G2a solid-phase sandwich enzyme-linked immunosorbent assay (ELISA) kit (#EH369RB; Invitrogen™ Human PLA2G2A ELISA Kit, Thermo Fisher, Carlsbad, CA) according to the manufacturer’s protocols. All samples were assayed in triplicate. Standard curves were used to derive PLA2G2A levels from best fit curves.

#### Lipopolysaccharide Binding Protein assay

Plasma samples were assayed for Lipopolysaccharide Binding Protein (LBP) by using a human LBP solid-phase sandwich enzyme-linked immunosorbent assay (ELISA) kit (#EH297RB; Invitrogen™ Human LBP ELISA kit, Thermofisher, Carlsbad, CA) according to the manufacturer’s protocols. All Samples were assayed in triplicate. Standard Curves were used to derive LBP levels from best fit curves.

### Breath Metabolite Analysis

#### Thermal desorption and GCxGC parameters

Prior to analysis, sorbent tubes were brought to room temperature and loaded into autosampler (Utra-xr, Markes International, UK). A gaseous standard mixture (1.01 ppm Bromochloromethane, 1.04 ppm 1,4-Difluorobenzene, 1.04 ppm Chlorobenzene-D5, 0.96 ppm 4-bromofluorobenzene) was immediately added to each tube, followed by a purge pre-desorption step consisting of 10 min with He at 50 mL.min^-1^, to remove water content in breath samples. Tubes were thermally desorbed for 10 min at 270°C (Unity-xr, Markes International, UK) and transferred to a “Universal” cold trap which matched the sorbent of the sample tube, held at 10°C and subsequently heated to 300°C, to minimize band broadening. The split flow after the cold trap was 15 mL.min^-1^.

Analysis by two-dimensional gas chromatography was conducted using an Agilent 7890B GC system, fitted with a flow modulator and a three-way splitter plate coupled to a flame ionization detector and a time-of-flight mass spectrometer with electron ionization (SepSolve, UK). Chromatographic analysis was performed using a Stabilwax (30 m × 250 μm ID × 0.25 μm df) as the first dimension (1D)-GC column and a Rtx-200 MS (5 m × 250 μm ID × 0.1 μm df) as second dimension (2D)-GC column, both purchased from Restek (Bellefonte, PA, US). The following GC oven temperature program was used: initial temperature 40°C was ramped to 215°C at 3°C.min^_1^ and held for 1 min. The oven temperature was then increased to 260°C at 50°C.min^_1^. The final temperature of 260°C was held for 10 min. The total run time for the analysis was 70 min. Helium carrier gas was flowed at a rate of 1.2 mL.min^_1^. The flow modulator (Insight, SepSolve Analytical, UK) had a loop with dimensions 0.53 mm i.d. x 110 mm length (loop volume: 25 uL), and the modulation time was 2 s total.

#### TOF-MS Conditions

The GCxGC was interfaced with a BenchTOF-select time-of-flight mass spectrometer (SepSolve Analytical, UK). The acquisition speed was 50 Hz and mass range was 35-350 m/z. The ion source and transfer line were set at 250 °C and 270 °C respectively and filament voltage at 1.6 V. Electron ionization energy was 70 eV. ChromSpace (SepSolve Analytical, UK) was used to synchronize and control the INSIGHT modulator, thermal desorption, GC, and TOF.

#### Chemical standards and solutions

Compounds shown in **Supplemental Table 2** were purchased from Sigma-Aldrich (St. Louis, MO, US). To spike the compound of interest into a sorbent tube, a 100 ppm solution was prepared in HPLC grade methanol. Using a solution loading rig (Markes International Limited, UK), 1 μL of the solution was spiked into a sorbent tube. The sorbent tube was flushed for 3 min with nitrogen at a flow of 100 mL.min^_1^. All the stock solutions were stored in glass vials and kept at 4°C. Sorbent tubes containing standards were analyzed by GCxGC BenchTOF-MS following the same protocols as described below for breath samples.

#### Quality control

To evaluate for changes in instrument sensitivity over time, a mixture of external standards was analyzed with the GCxGC BenchTOF-MS alongside the breath samples, as described previously^27^. Briefly, we analyzed an external standard before running each batch of breath samples. The standard used was EPA 8240B Calibration Mix (2-butanone, isobutanol, 4-methyl-2-pentanone and 2-hexanone). One mL 2000 µg.mL^-1^ vial standards were purchased from Sigma-Aldrich. To spike the mixture into a sorbent tube, a 100 µg.mL^-1^ solution was prepared in HPLC grade methanol. Using a solution loading rig (Markes International Limited, UK), 1 μL of the solution was spiked into a sorbent tube. The sorbent tube was flushed for 3 min with nitrogen at a flow of 100 mL.min^-1^ and analyzed by GCxGC BenchTOF-MS.

#### Untargeted data analysis

GCxGC–MS deriving chromatograms were first aligned to one user-selected reference chromatogram in ChromCompare+ software version 2.1.4 (SepSolve Analytical Ltd, UK) based on the 1D and 2D retention times and the available spectral information. The alignment algorithm was used to overcome retention time drift observed across the dataset.

A tile-based approach was applied to the aligned chromatograms to enable the raw data to be imported into the chemometrics platform directly, without the application of any pre-processing methods, such as integration and identification. The tile size was 32 s in 1D and 0.5 s in 2D with 25% overlap. The signal for every individual m/z channel of each tile was integrated for comparison across every chromatogram in the dataset. This process generated a list of features labeled according to the tile retention times and m/z channel.

The data matrix was cleaned of possible artifacts and siloxane derived from the sorbent, and filtered so that individual m/z channels with < 25000 intensity were removed. A feature was retained if it was present in more than 50% of the samples in either group (i.e. MIS-C or non-MIS-C), and data were then normalized using internal standard (4-bromofluorobenzene).

Data reduction was performed using the proprietary feature selection algorithm in ChromCompare+ software. The algorithm uses a multivariate method to consider the covariance between features^45^; this approach retained the top 50 most significant features of the known sample classes (i.e. MIS-C versus non-MIS-C). Of these features, 29 features resulted in unique volatiles.

Unique peaks were putatively identified via comparison with the NIST20 library based on the combination of the mass spectra similarity match ≥60% and further identity confirmation was obtained with pure standards.

Data visualization (heatmaps, box plots and ROC curve) were performed using RStudio v1.3.1073 (PBC, Boston, MA) and GraphPad Prism V.8.4.3 (GraphPad Software, San Diego, CA).

### Machine Learning Methods

#### Data pre-processing

We pre-processed the clinical features data, lab measurements, cytokines (SC5b9 and PLA2G2A concentration), flow cytometry, and breath volatiles. **Figure 1c** illustrates the sample overlap between the various data measurements for the cohort. For cytokines (SC5b9 and PLA2G2A concentration) and lab measurements, we feature engineered to account for the concentrations below limit of detection (LOD) threshold. For the classification model, we built individual models for each data measurement and an integrated model. Because inclusion of breath volatiles measurements to the integrated model decreased the power of our analysis (due to small sample sizes), these were removed from the integrated models. To assess the collinearity between the features within data measurement and across data measurements, we performed Pearson correlation (**Supplemental Table 1**). Features that were highly correlated (p-value <0.5) were excluded from the model analysis. All the computational analysis was performed using Python (*sklearn, pandas*).

#### Random forest with stratified k-fold cross-validation

In order to classify a patient’s MIS-C status, we built a random forest classification model yl7J(X) with stratified k-fold cross validation on a full set of data, {LL,LL} LL=1. We fit the model on full dataset due to small sample size. Each of the N examples represents a patient where LLLL refer to a matrix of predictors/features, with N rows and d columns and LL{0,1} is the patient’s outcome, encoded as 1 if the patient is diagnosed with MIS-C and 0 otherwise. Random Forest Classifier is an ensemble machine learning model, combines multiple decision trees using bootstrap aggregating and random feature selection. The model’s output is determined by a majority vote from individual trees. Notably, it can estimate feature importance using Gini Importance, calculated by the mean decrease in the heuristic resulting from splitting on a feature across the forest. Normalization ensures that all Gini importance scores sum to 1.

To address the challenge of a small sample size, we performed an analysis on the full dataset, generating a Receiver Operating Characteristic (ROC) curve and a confusion matrix. Given the class imbalances in the dataset, we employed stratified k-fold cross-validation, which ensures that the class proportions are maintained in each fold. This approach prevents over-representation of one class, especially crucial when dealing with unbalanced datasets. We predicted the actual class and class probability (i.e., the probability of the instance to be classified into class 0 or 1) for the whole dataset. The confusion matrix was plotted for various predicted probability thresholds. We explored 6 thresholds (0.25, 0.30, 0.35, 0.40, 0.45, 0.48) for prediction and chose the threshold to strike a balance between type I and type II errors.

