## Supplemental Table 2 and 3 for "Metabolomic and Immunologic Discriminators of MIS-C at Emergency Room Presentation"

**Short title: MIS-C diagnosis**

**Authors:**

Laura A. Vella^1,2^*^#^, Amalia Z. Berna^1^*, Allison M. Blatz^1,3^, Joey Logan^4^, Priya Sharma^4^, Yang Liu^1^, Jonathan Tedesco^1^, Cara Toland^1^, Leena Babiker^1^, Kathryn Hafertepe^1^, Shane Kammerman^1^, Josef Novacek^1^, Elikplim Akaho^1^, Alexander K. Gonzalez^4^, Deanne Taylor^2,4^, Caroline Diorio^2,5^, Fran Balamuth^2,6^, Hamid Bassiri^1,2^, Audrey R. Odom John^1,2#^

**Affiliations:**

^1^Division of Infectious Diseases, Children's Hospital of Philadelphia, Philadelphia, Pennsylvania, USA

^2^Perelman School of Medicine, University of Pennsylvania, Philadelphia, Pennsylvania, USA

^3^Nemours Children’s Health, Wilmington, Delaware, USA

^4^Department of Biomedical and Health Informatics, Children's Hospital of Philadelphia, Philadelphia, Pennsylvania, USA

^5^Division of Hematology and Oncology, Children's Hospital of Philadelphia, Philadelphia, Pennsylvania, USA

^6^Division of Emergency Medicine, Children's Hospital of Philadelphia, Philadelphia, Pennsylvania, USA

***co-first authors**

**^#^Correspondence:**

Audrey R. Odom John MD PhD

Children’s Hospital of Philadelphia

3501 Civic Center Blvd

CTRB 10100

Philadelphia PA 19104-4318

and

Laura A. Vela MD PhD

Children’s Hospital of Philadelphia

3501 Civic Center Blvd

CTRB 10008

Philadelphia PA 19104-4318

**Supplemental Table 2:** List of top discriminatory compounds between MS-C and non-MIS-C febrile patients, together with analytical characteristics of each compound

| Compound Name | Formula | Structure | Rt_1_  min | Rt_2_  sec |
| --- | --- | --- | --- | --- |
| Acetone | C3H6O | 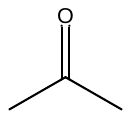 | 8.267 | 2.125 |
| Ethyl acetate | C4H8O2 | 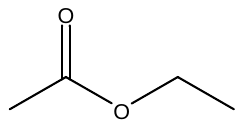 | 9.067 | 0.625 |
| Nonane, 2-methyl- | C10H22 | 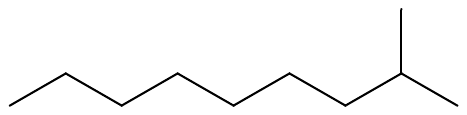 | 10.267 | 0.625 |
| Unknown1 |  |  | 11.467 | 0.625 |
| Cyclohexene, 4-ethenyl- | C8H12 | 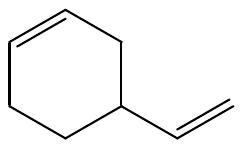 | 11.867 | 0.625 |
| α-Pinene | C10H16 | 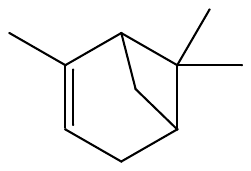 | 12.267 | 0.625 |
| Camphene | C10H16 | 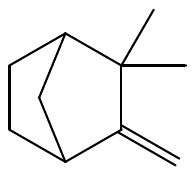 | 14.267 | 0.625 |
| β-pinene | C10H16 | 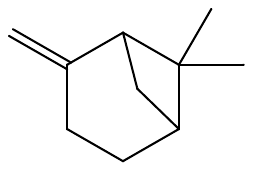 | 15.467 | 0.625 |
| β-Myrcene | C10H16 | 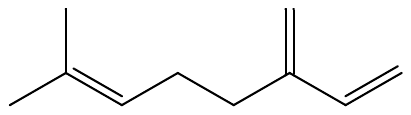 | 17.467 | 0.25 |
| Cumene | C9H12 | 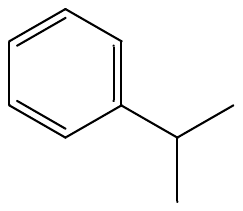 | 18.267 | 0.625 |
| Dodecane | C12H26 | 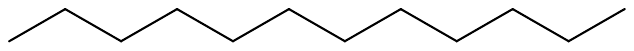 | 18.667 | 1.00 |
| D-Limonene | C10H16 | 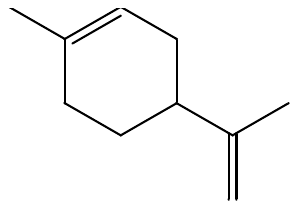 | 19.067 | 0.25 |
| Carveol | C10H16O | 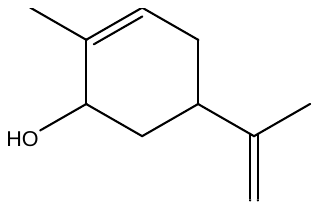 | 20.267 | 0.25 |
| Eucalyptol | C10H16O | 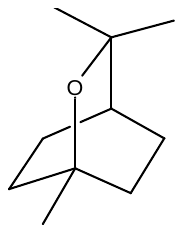 | 20.267 | 0.625 |
| γ-Terpinene | C10H16 | 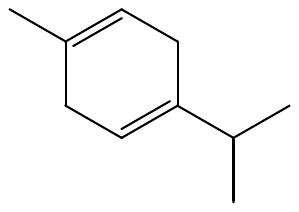 | 21.067 | 0.25 |
| p-Cymene | C10H14 | 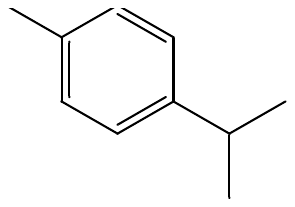 | 22.267 | 0.25 |
| Terpinolene | C10H16 | 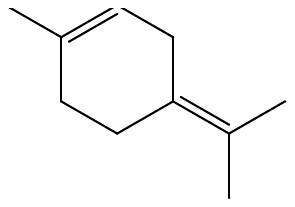 | 22.667 | 0.25 |
| ψ-Cumene | C9H12 | 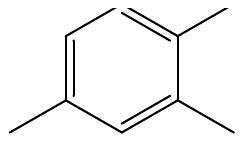 | 23.067 | 0.625 |
| 3-Hexen-1-ol, (Z)- | C6H12O | 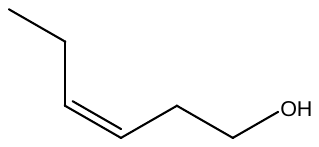 | 27.067 | 0.25 |
| 2-Ethyl-1-hexanol acetate | C10H20O2 | 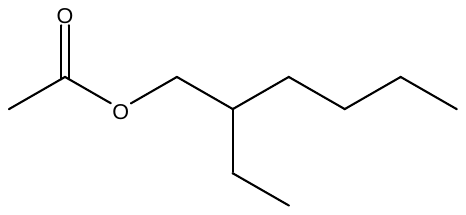 | 27.467 | 0.625 |
| Cyclohexanol | C6H12O | 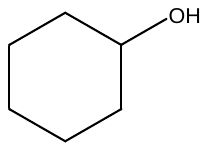 | 28.267 | 0.25 |
| p-Cymenene | C10H12 | 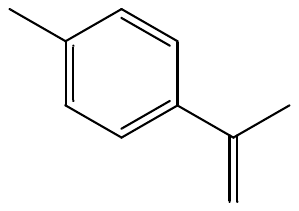 | 29.867 | 0.25 |
| 2-Ethyl-1-hexanol | C8H18O | 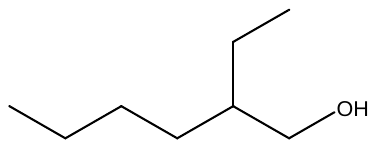 | 31.867 | 0.25 |
| Linalool | C10H18O | 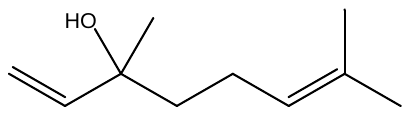 | 33.867 | 0.25 |
| Unknown2 |  |  | 35.867 | 0.25 |
| Levomenthol | C10H20O | 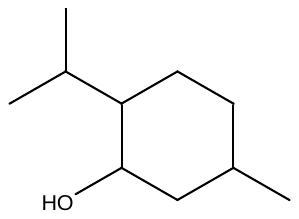 | 36.667 | 0.25 |
| Unknown3 |  |  | 39.867 | 0.25 |
| Unknown4 |  |  | 40.267 | 0.25 |
| Unknown5 |  |  | 41.067 | 2.125 |

**Supplemental Table 3:** List of flow cytometric antibodies. Panel is listed as run for reproducibility. Not all components were included in the features submitted for machine learning.

| **Protein** | **Clone** | **Fluorophore** | **Company** |
| --- | --- | --- | --- |
| CD45RA | HI100 | BUV395 | BD Biosciences |
| CD8 | RPA-T8 | BUV496 | BD Biosciences |
| CD19 | SJ25C1 | BUV563 | BD Biosciences |
| CD16 | 3G8 | BUV615 | BD Biosciences |
| CD38 | HIT2 | BUV661 | BD Biosciences |
| CD27 | L128 | BUV737 | BD Biosciences |
| CD20 | SH7 | BUV805 | BD Biosciences |
| SPIKE | - | BV421 Streptavidin | Spike (R&D Systems #AVI10586-050)  Biolegend |
| CD138 | MI15 | Pacific Blue | Biolegend |
| IgD | iA6-2 | BV480 | BD Biosciences |
| CD3 | UCHT1 | BV570 | Biolegend |
| CD39 | A1 | BV605 | Biolegend |
| CCR7 | G043H7 | BV650 | Biolegend |
| CX3CR1 | 2A9-1 | BV711 | Biolegend |
| CD4 | SK3 | BV750 | BD Biosciences |
| HLA-DR | G46-6 | BV786 | BD Biosciences |
| TCRβ21.3 | REA894 | FITC | Miltenyi Biotec |
| PD-1 | EH12.1 | BB700 | BD Biosciences |
| IgG | IS11-3B2.2.3 | PerCP-Vio700 | Miltenyi Biotec |
| DECOY | - | BB790 Streptavidin | BD Biosciences  Custom Conjugate Model: 624296-2282-1722-X-X-X |
| NUCLEOCAPSID | - | PE Streptavidin | Nucleocapsid (R&D Systems)  Biolegend |
| IgA | REA1014 | PE-Vio615 | Miltenyi Biotec |
| EOMES | WD1928 | PE-eFluor610 | Invitrogen |
| ICOS | C398.4A | PE-Cy5 | Invitrogen |
| T-BET | 4B10 | PE-Cy7 | Biolegend |
| RBD | - | APC Streptavidin | RBD (R&D Systems)  Biolegend |
| Ki-67 | B56 | AlexaFluor700 | BD Biosciences |
| VIABILITY DYE | - | Zombie NIR | Biolegend |
